# 3d virtual histology reveals pathological alterations of cerebellar granule cells in multiple sclerosis

**DOI:** 10.1101/2022.10.07.22280811

**Authors:** Jakob Frost, Bernhard Schmitzer, Mareike Töpperwien, Marina Eckermann, Jonas Franz, Christine Stadelmann, Tim Salditt

**Affiliations:** Institut für Röntgenphysik, Georg-August-Universität Göttingen, Germany; Institute of Computer Science, Georg-August-Universität Göttingen, Germany; Institut für Neuropathologie, Universitätsmedizin Göttingen, Germany; Cluster of Excellence ‘Multiscale Bioimaging: from Molecular Machines to Networks of Excitable Cells’ (MBExC), Georg-August-Universität Göttingen, Germany; (M.E.) ESRF, Grenoble, France; (M.T.) XYLON GmbH, Hamburg, Germany

## Abstract

We investigate structural properties of neurons in the granular layer of human cerebellum with respect to their involvement in multiple sclerosis (MS). To this end we analyze data recorded by X-ray phase contrast tomography from tissue samples collected post mortem from a MS and a healthy control group. Using automated segmentation and histogram analysis based on optimal transport theory (OT) we find that the distributions representing nuclear structure in the granular layer move to a more compact nuclear state, i.e. smaller, denser and more heterogeneous nuclei in MS. We have previously made a similar observation for neurons of the dentate gyrus in Alzheimer’s disease, suggesting that more compact structure of neuronal nuclei which we attributed to increased levels of heterochromatin, may possibly represent a more general phenomenon of cellular senescence associated with neurodegeneration.

## Introduction

The complex cytoarchitecture of the human brain can undergo pathological alterations associated with neurodegenerative diseases. Morphological changes may range from drastic and relatively easy to be diagnosed to – on the contrary – very subtle and yet elusive changes. Deciphering the interplay between the neuronal tissue structure and the development of neurodegenerative diseases therefore remains a challenge which requires further progress in imaging and morphometric quantification. Today, histology and histopathology is largely based on tissue sections and observation of exemplary regions in two-dimensions (2d) by optical or electron microscopy. Three-dimensional (3d) imaging is required to digitalize and compare structures in their full dimensionality. Serial sectioning, staining, digital microscopy and subsequent alignment is laborsome, but can in principle address micro-anatomy and cytoarchitecture in 3d. Unfortunately, this comes at the cost of a non-isotropic resolution, possible artifacts due to the slicing, staining, or the alignment procedure. Moreover, this approach is severely limited in throughput, and therefore often impedes the visualization of large fields of view (FOV), even at moderate resolution, as well as the comparison between a sufficient number of individuals. Sufficient sample size and volume, in combination with unsupervised morphometric quantification, however, is a prerequisite to understand the limits of ‘structural homeostasis’ and the onset of pathological structural alterations. X-ray phase contrast computed tomography (XPCT) has been recently introduced as a new 3d imaging method for histology and pathohistology ***Albers et al. (2018***); ***Massimi et al. (2020***); ***Vågberg et al. (2018***); ***Reichardt et al. (2021***); ***Frohn et al. (2020***); ***Dejea et al. (2019***); ***Khimchenko et al. (2016***). It offers a capability for high resolution imaging of soft tissues over a cross section of several mm, with a geometric zoom able to visualize selected regions of interest down to 20 nm to 50 nm voxel sizes ***Bosch et al. (2022***); ***Kuan et al. (2020***). In this way, 3d reconstructions of neurons and their spatial organization within particular regions can be obtained. By comparison of different cohorts, morphometrical analysis may also contribute to an understanding of neurodegenerative mechanisms, in particular if tissue structure is probed at subcellular resolution.

A case in point is cerebellar involvement in multiple sclerosis (MS), which is relevant for MS-related impairments, including not only cerebellar motor dysfunction but also cognitive cerebellum associated deficits ***Weier et al. (2015***); ***Kutzelnigg et al. (2007***); ***Parmar et al. (2018***). More generally, as MS research is no longer restricted to inflammation and demyelination as major pathological mechanisms, but also includes neurodegenerative processes ***Albert et al. (2017***), it is timely to address the cerebellar cytoarchitecture in a broader sense. As one of the oldest brain regions of mammals, the cerebellum is well known for its role in motion control and synergy of movements ***Manto et al. (2013***). Despite a relatively small weight of ∼ 10% of the total brain mass, the cerebellum contains 80% of the total number of neurons within the human brain ***Azevedo et al. (2009***). Tightly packed neurons in its densest layer, the so-called granular layer, are a particular target when screening for possible MS-related changes in the cytoarchitecture. Along with the molecular layer and the interfacial Purkinje cell layer, the granular layer is part of the tightly folded cerebellar cortex.

In a preceding study, we provided 3d imaging of tissue samples from human cerebellum, collected post mortem by autopsy ***Töpperwien et al. (2018***). The millimeter sized samples were taken by biopsy punches from formalin-fixed and paraffin-embedded (FFPE) tissue blocks, and scanned by XPCT. Phase contrast was achieved based on propagation of partially coherent wavefields, using both synchrotron radiation for high resolution and custom *µ*-CT scanners for larger overviews. Datasets with subcellular resolution were obtained, and a reconstruction workflow to automatically locate the neuronal nuclei in the molecular and granular layer by an automated approach based on the Hough transform, giving a detailed statistical account of the spatial packing of neurons within the granular layer (cf. Fig. 1d). Based on local density estimations and pair correlation functions, a previously unknown anisotropy in the short-range order of granule cells was reported, which reflected the plane of the dendritic trees of the Purkinje cells.

**Figure 1.**
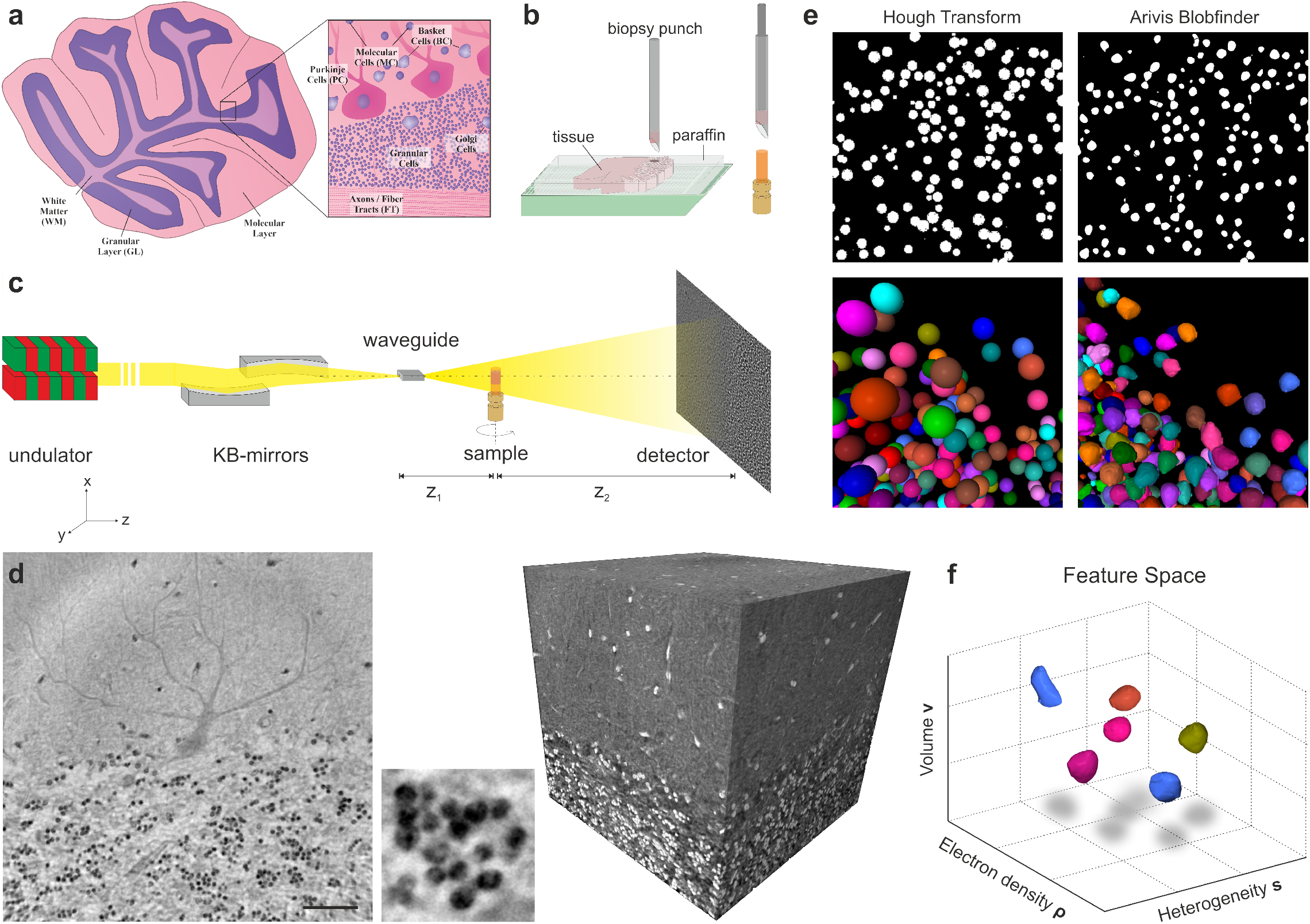
Experimental setup and analysis workflow. (**a**) Sketch of the human cerebellum in transversal slice and zoom-in to the cerebellar cortex, which contains various types of cells. (**b**) Tissue samples of the cerebellum were taken post mortem from twelve individuals (6 MS, 6 Control) and embedded in paraffin. Biopsy punches of the samples were placed into a Kapton tube for scanning. (**c**) Schematic of the synchrotron setup at the PETRAIII storage ring (DESY, Hamburg). X-rays with an energy of 13.8 keV are generated, focused and hit an intensity detector behind the sample. From the projections, the sample is reconstructed using phase retrieval. (**d**) The reconstructed volume covers the interface between the molecular- and the granular layer (right, inverted contrast). A virtual slice through the volume reveals histological features such as the granule cell nuclei or the dendritic tree of a Purkinje cell (left). Scale bar: 50 µm. Also the internal structure of the nuclei can be resolved (middle). (**e**) In the previous work of ***Töpperwien et al. (2018***) the data were segmented with the Hough transform, which is suited to find center positions of the GC-nuclei and generates spherical objects (left). Here, we segment the nuclei with the Blob Finder algorithm from Arivis, which generates segments covering the actual shape of the nuclei (right). This enables to determine structural features of the nuclei such as their volume or sphericity. Note that the 3d views in (e) left/right do not correspond to the same location. (**f**) The nuclei are then represented in a feature space – an abstract space in which each nucleus represents a point with coordinates encoding their structural properties. Each subject is represented by a point cloud, with is then further analyzed in view of pathological alterations.

In this work, we extend the previous analysis from physiological histology to the pathohistology of MS. To this end, we use the data provided in ***Töpperwien et al. (2018***) and investigate possible alterations in the granular layer occurring in the tissue of 6 MS patients compared to samples from 6 control subjects. Since data acquisition and tomographic reconstruction was already reported in detail ***Töpperwien et al. (2018***); ***Töpperwien (2018***), we concentrate here on pathological structural alterations of cerebellar granule cells. Note that previously only neuron locations, but no structural features of neurons were obtained in the segmentation. Here we can now provide a detailed comparison of structural features of neuronal nuclei, since progress in segmentation allowed us to extract not only positions of nuclei, but also size, shape, and electron density, as well as heterogeneity of the density within the nucleus. Furthermore, we now have novel statistical tools at hand, based on optimal transport (OT) theory ***Santambrogio (2015***); ***Peyré and Cuturi (2019***), with which the structural differences in the cytoarchitecture can be compared, even without prior structural hypothesis and group attribution. The fact that we earlier found clear changes in dentate gyrus granule cells associated with Alzheimer’s disease (AD) ***Eckermann et al. (2021***), also motivated us to reinvestigate the cerebellum data, in view of possible involvement of cerebellar granule cells in MS.

The manuscript is organized as follows: after this introduction, the results section first describes the available data and the segmentation of neuronal nuclei, before we analyze neuronal density and packing, and then nuclear morphological features. The multidimensional histograms representing the neuron population of any single subject are then further compared between individuals of the MS and control groups, using concepts of OT theory. The paper closes with a discussion and summary of the main morphometric results. Technical details are presented in the Materials and Methods section in summarized form, and in part further explained in the appendix.

## Results

Figure 1 presents (a) a schematic of the granular layer as the target region of this work, and (b) the sample extraction from a tissue block, which is chosen to probe the granular layer of the cerebellar cortex, consisting of the molecular layer, the granular layer and the Purkinje cell layer at the interface of the latter. In (c), the XPCT data acquisition scheme is depicted, followed by (d) an example illustrating the data quality, (e) a comparison of two different segmentation methods, and finally (f) a schematic of the so-called feature space consisting of point clouds, in which the coordinates of each point represents features of corresponding nuclei, which then subjected to further analysis.

### Data

Tissue samples were acquired from a total of twelve age-matched subjects: six patients suffering from multiple sclerosis (MS), and six healthy control subjects (Control, CTRL). The samples were collected post mortem, chemically fixed and embedded in paraffin. Using biopsy needles, cylindrical tissue samples were punched out of the paraffin-embedded tissue blocks, and placed in a polyimide (Kapton) tube, as depicted in Fig. 1(b). The samples were scanned at the GINIX endstation ***Salditt et al. (2015***) of the P10 beamline of the PETRAIII storage ring (DESY, Hamburg), see the schematic of the experimental setup in Fig. 1(c). The monochromatic (Si(111) channel-cut monochromator) undulator beam of 13.8 keV photon energy was prefocused by a pair of Kirkpatrick-Baez (KB) mirrors and coupled into an X-ray waveguide serving further spot size reduction and coherence filtering. Using this scheme, the samples which are positioned on the fully motorized tomographic stage are illuminated by a fully coherent beam with reduced wavefront artefacts which facilitates a clean and artifact poor image formation. Projected in-line holograms are recorded on a fibre-coupled detector. Projection images are first treated by phase retrieval using a Contrast-transfer function (CTF)-based algorithm ***Cloetens et al. (1999***) implemented in a published software package ***Lohse et al. (2020***) prior to tomographic reconstruction. Detailed information about data acquisition and the experimental setup can be found in ***Töpperwien et al. (2018***). The reconstructed samples have a field of view (FOV) of 336 × 336 × 375 µm^3^ and capture the transition from the molecular to the granular layer. The voxel size of 187 nm is sufficient to identify various histological characteristics such as blood vessels, the dendritic tree of a Purkinje cell as well as the nuclei of the granule cells, when observing a virtual slice through the volume (see e.g. Fig.1(d) left). Furthermore, the internal structure of the nuclei is resolved. Note that gray values in XPCT represent phase shifts of the X-ray beam which are proportional to the electron density difference with respect to the average sample, here predominantly the paraffin mounting medium. In ***Töpperwien et al. (2018***) the granule cell nuclei were already segmented using the spherical Hough transform ***Peng et al. (2007***). The Hough transform finds the center positions of spherical objects and generates spheres of equal size. Contrarily, here we are interested in the details of nuclear volumes, shapes, and electron density distribution. To this end, the nuclei are segmented with the Blob Finder algorithm of the software Arivis (Zeiss AG, Germany). The Blob Finder generates segments which cover the actual shape of the nuclei, in contrast to the Hough transform which only gives the center positions of the nuclei. Here we can hence exploit detail structural properties related to the size, roundness and the density of the nuclei, as illustrated in Fig.1(e). We use this information to place each neuron in a *feature space*, with coordinates representing the corresponding structural property, as schematically illustrated in Fig.1(f) and previously introduced in ***Eckermann et al. (2021***). In this way, one obtains a multidimensional distribution (histogram) for each subject. Analysis of the entire distributions and OT based metrics then allows us to probe differences between individuals on the histogram level, instead of only the mean or median values. Most notably, we can compare data beyond the capacity limits and bias of visual inspection.

### Segmentation

The segmentation was performed using the software Arivis Vision4D (Zeiss AG, Germany). Several ten thousand neurons were detected in a semiautomatic workflow for each sample. The segments were created with the Blob Finder algorithm which is designed to find round, sphere-like objects. It has several adjustable parameters with which size and number of found objects can be varied. Note that all samples were segmented with the same parameters in order to make the results comparable. Since we focus on the analysis of the granule cell nuclei, all objects found by the Blob Finder outside the granular layer were vetoed out. For this purpose, a mask was created which only encloses the granular layer. The mask was defined by manual drawing in the data, see Fig.2(a). All objects outside the mask are removed. Furthermore, several filters were applied to remove segments which clearly do not represent granule cell nuclei. For detailed information about the filter limits and the general segmentation workflow, see Appendix 1. The limits of the filters were determined by plotting all segments in a scatter plot, see Fig. 2(b). In the scatter plot, all segments are plotted as single points according to their properties like volume or sphericity. In this representation, three subgroups of segments can be identified. The middle of the three groups are identified as the granule cells. The left group are small artifact segments which are filtered out by a volume filter of ≈ 10 µm^3^. The right group corresponds to “double cells”, which occur when two cells are close to each other and are covered by only one segment. To ensure that each cell is covered by exactly one segment, the double cell segments can be separated with the splitting operation of Arivis. This operation applies a distance map on the segments, whose local maxima are then used as seed points for a Watershed-algorithm. Figure 2(c,d) show the final segmentation results after filtering. A homogeneous distribution of round segments is obtained, which adequately covers the granule cell nuclei. From the segments the following properties are extracted: the center of mass, volume, sphericity, mean of electron density and standard deviation of electron density. The latter is dominated not by white shot noise of the reconstructed gray values, but by the density variations representing nuclear sub-structure. We denote this variation as *heterogeneity* of the nucleus.

**Figure 2.**
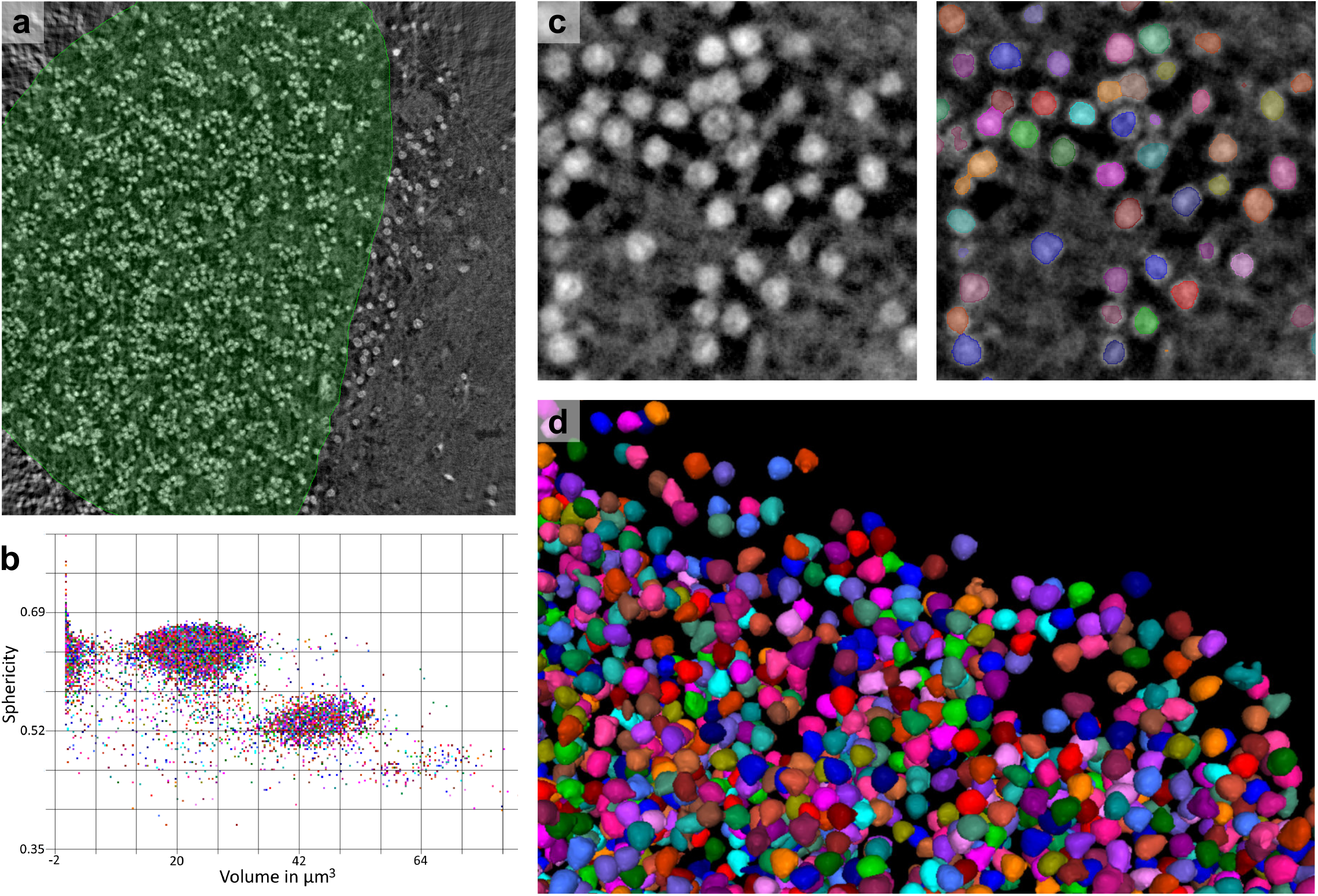
Segmentation of the granule cell nuclei. (**a**) A mask enclosing the granule layer is used to remove segments lying in the molecular layer and Purkinje cell layer as well as artifacts in the corner of the samples. (**b**) Plotting the properties volume and sphericity of all segments in a scatterplot allows the segments to be distinguished into three subgroups: granule cells (middle), double cells (bottom right) and artifacts (left). The artifacts are filtered out, and the double cell segments are split into single cell segments (**c**) The final segmentation results shown in a 2d slice. Different segment colors serve for better distinction. (**d**) Three-dimensional rendering of the segments reveals a homogeneous distribution of round spheres, which adequately represent the granule cell nuclei.

### Density and local ordering of granule cells

With the center positions of the nuclei obtained from the segmentation, the density of the granule cells within the granular layer is calculated for each subject. To this end, the volume of the granule layer is computed by creating an envelope around all cells. The total cell count divided gives the average cell density, which is plotted for all subjects in Fig. 3c. For the MS group, the average 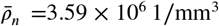 is higher than for Control subjects with 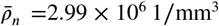, with marginal statistical significance of *p* = 0.091 (Welch’s t-test, double-sided). The short range order of the cells can be further investigated by calculating the structure factor *S*(**q**) for each sample, using concepts of quantifying ordering in amorphous and liquid structures of condensed matter, which we had already used for neurons in ***Töpperwien et al. (2018***). The structure factor is calculated based on the nuclei center positions **r** according to

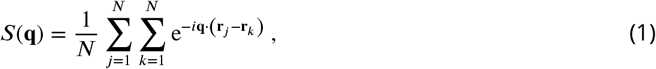

where *N* denotes the number of nuclei and **q** the reciprocal space vector. A 3d structure factor *S*(**q**) is obtained for each subject, which was investigated in view of directional anisotropy already in ***Töpperwien et al. (2018***). Here, we focus on the comparison between MS and CTRL and to this end content ourselves with the 1d (powder averaged) *S*(*q*), which can be compared more easily already based on visual inspection. Figure 3a shows the group-averaged *S*(*q*) curves, representing a mean structure factor for MS and one for CTRL. Two strong modulations of the curves show that the neurons exhibit pronounced short range order. The peak of the MS curve is slightly shifted towards the higher wave numbers in comparison to *S*_CTRL_(*q*), which indicates a more compact arrangement in MS and is in agreement with the higher cell densities. To determine whether the observed shift is statistically significant, we test whether the graphs lie within each other’s error interval. As error interval, the standard error of the mean 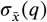 *is chose*n. At every point *q* where the structure factors are sampled, the squared ratio *χ*^2^ of the distance between the graphs and the error 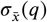 is calculated and averaged over all *q* as

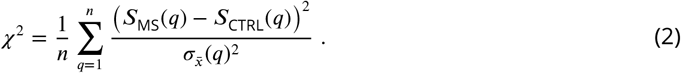

**Figure 3.**
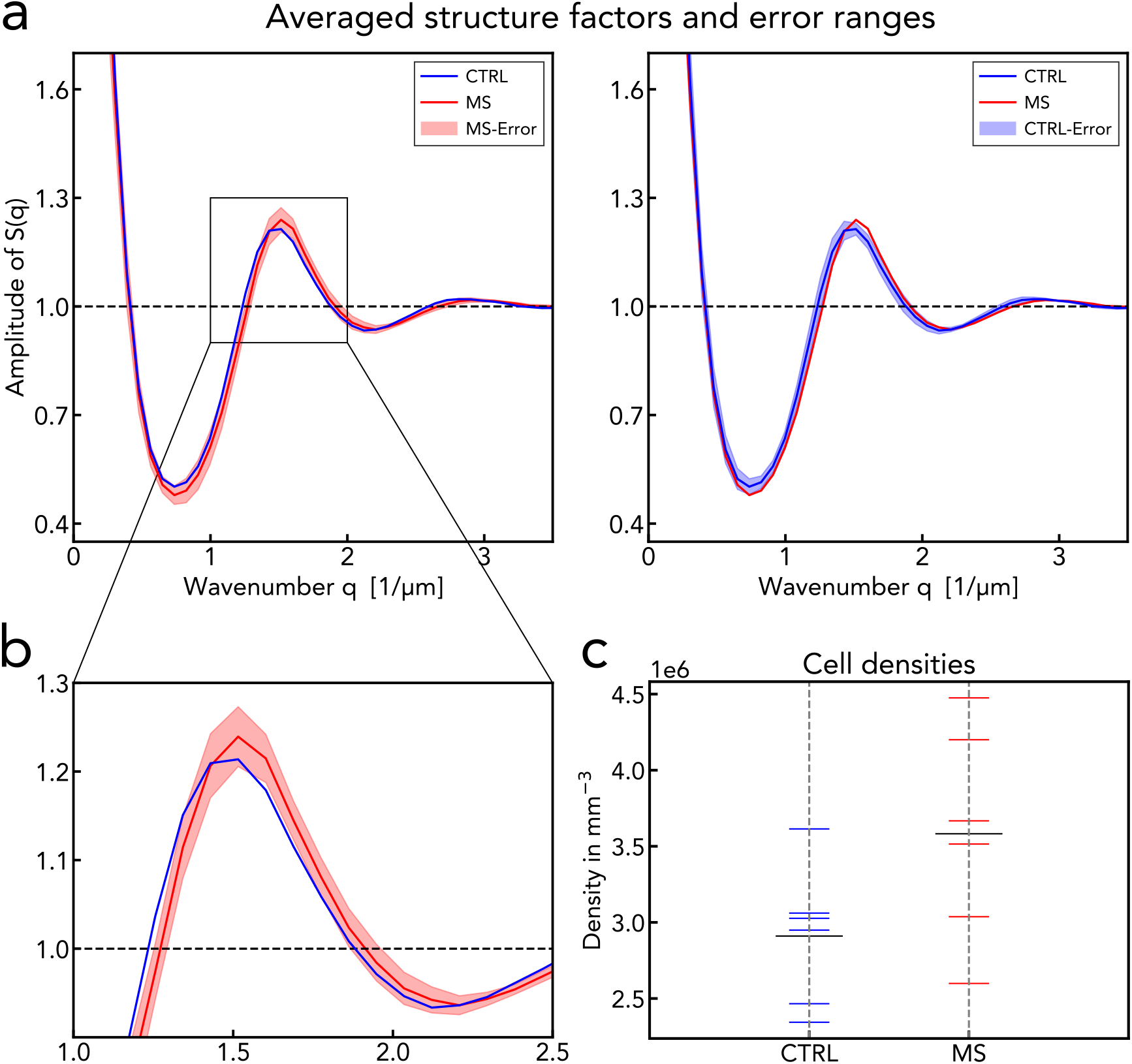
Structure factor characterizing spatial arrangement of the nuclei. (**a**) The group averages of the powder-averaged structure factor, with the error intervals (standard error of the mean 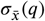) (left) of the MS-group (left) and (right) the CTRL-group. (**b**) A zoom-in showing the MS group-averaged structure factor to be located at the boundary of the error interval of the CTRL group, and vice versa, which is quantified by *χ* values of 1.04 and 1.58, respectively. (**c**) The cell density of all individuals plotted with the mean density of both groups. t-test on the density values yields *p* = 0.091.

Since the powder-averaged structure factors are noisy for very high and very small *q* values, this is done only in the range 0.3 µm < *q* < 5 µm (5 µm is the sampling limit of the 3d structure factors). Note that two *χ*^2^ values are obtained, since we can compare the distance once with the error of MS and once with that of CTRL, resulting in *χ*^2^ = 1.04 and *χ*^2^ = 1.58 respectively. This would indicate that the differences are not statistically significant, and that more samples are required to unravel possible inter-group effects from the inter-subject variance. In fact, when inspecting the residuals and the systematic changes in the curve, one may very well be tempted to reject the null hypothesis.

### Creating the feature space

After the granule cell nuclei are segmented and several properties have been extracted, we next investigate whether the structural properties of the nuclei (as opposed to their spatial arrangement and ordering treated above) exhibit significant systematic changes associated with MS pathology. To this end, we have created a workflow in which the nuclei are characterized by several quantifiable properties, which we denote as features. Accordingly, each segmented nucleus can be considered as a point in a so-called “feature space”. The coordinates of the point are given by the respective feature values, where each dimension corresponds to one feature. For every subject, the population of all granule nuclei then forms a point cloud in this feature space. The point cloud can be thought as a sum of Dirac masses with uniform weights 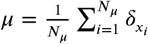. Thus, one receives a multidimensional discrete distribution, which represents an individual by the properties of its nuclei. The following six features were chosen for the analysis:

- volume *ν*
- sphericity *φ*
- mean of electron density *ρ*
- heterogeneity *s*
- number of neighbors within local vicinity *nn*
- distance to nearest neighbor *d*_*nn*_

The heterogeneity describes the standard deviation of the electron density within the nucleus, given by 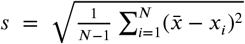 where *x*_i_ are the gray values (≙ electron density) of the voxels enclosed by a segment. The number of neighbors *nn* is given by the number of cells located within a radius of 7.45 µm from the original cell. The value of 7.45 µm was chosen such that it corresponds to the average position of the local minimum between the first and second correlation shell of the pair correlation function. Prior to analysis, the data is standardized and normalized. To do so, the mean value *µ* of the total population, that is all granule cells of all subjects, is subtracted from each individual measurement *x* and divided by the standard deviation *σ* of the population with *z* = (*x* − *µ*)/*σ*. This is done separately for each feature. The standardized population then has an expectation value of zero and a variance of one. After standardization, we construct a 6d feature space out of the six features. Figure 4a shows 2d projections of a point cloud in feature space. Plotting the individual features of all nuclei from a subject as histograms (see Fig. 4b), shows that all features are approximately Gaussian distributed. We first examine differences between groups by creating violin plots out of the 1d histograms. When doing this for the heterogeneity *s*, a clear trend between groups can be identified (see Figure 4c). The median values of the MS group are all higher than those of the Control group. However for the remaining features, no distinct trends can be found (cf. Appendix 3). Furthermore, the median values over the neuron populations for all features are calculated. Figure 4d shows the median values of all individuals and for each feature. Again, a significant difference between the groups can be found for the heterogeneity *s*, whereas for the remaining features, no statistically significant result can be inferred. In order to quantify group differences, a t-test (Welch, double-sided) is applied to the median values. The calculated p-values are listed together with the averaged median values of the nuclei parameters in Table 1. A *p* value of 0.001 confirms a significant difference in heterogeneity, whereas for the other features no differences were found between Controls and MS patients. Note that the *p* values reflect group differences with respect to the twelve median values rather than to the entire neuron population, which we investigate next in more detail.

**Table 1.** Overview of granule cell parameters. The data and *p* values are calculated from the median values of the subject populations prior to standardization. The *p* value is with 0.001 very low for heterogeneity. The values of the electron density indicate the difference to the electron density of the average medium, which is paraffin in this case. *Since *nn* takes only integer values, the population mean instead of the median was used for reasons of accuracy.

| Median over subject<br>neuron population | ALL | | CTRL | | MS | | $p^*$ |
| --- | --- | --- | --- | --- | --- | --- | --- |
|  | mean | std | mean | std | mean | std |  |
| Rel. electron density $\rho$ (1/nm<sup>3</sup>) | 31.23 | 5.38 | 29.19 | 5.02 | 33.26 | 4.85 | 0.225 |
| Heterogeneity $s$ (1/nm<sup>3</sup>) | 8.78 | 0.94 | 8.01 | 0.56 | 9.55 | 0.51 | 0.001 |
| Volume $v$ (μm<sup>3</sup>) | 26.83 | 4.41 | 28.35 | 5.35 | 25.3 | 2.38 | 0.283 |
| Sphericity $\phi$ | 0.62 | 0.01 | 0.63 | 0.01 | 0.62 | 0.01 | 0.569 |
| $nn$ within radius of 7.45 μm* | 7.24 | 1.2 | 6.83 | 0.75 | 7.66 | 1.41 | 0.281 |
| Nearest-neigh. dist. $d_{nn}$ (μm) | 4.38 | 0.22 | 4.44 | 0.2 | 4.32 | 0.22 | 0.391 |
| Cell density $\bar{\rho}_n$ (·10<sup>6</sup>/mm<sup>3</sup>) | 3.05 | 0.64 | 2.99 | 0.43 | 3.61 | 0.64 | 0.091 |

**Figure 4.**
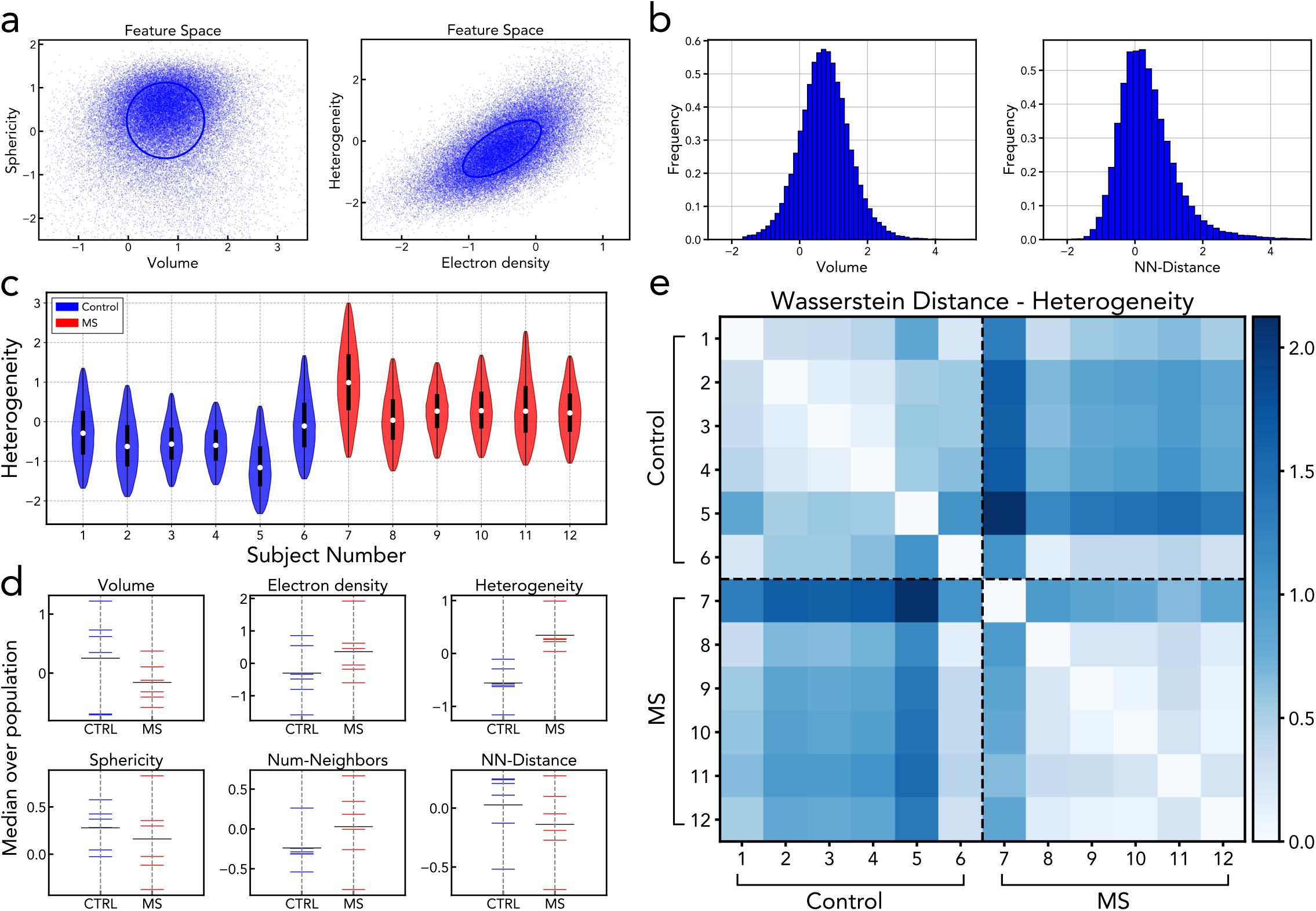
Features space and individual feature analysis. (**a**) From the segmented nuclei, a feature space is constructed in which individuals are represented by the features of their nuclei. The figure shows 2d projections of the 6d feature space, example shown for one subject. Single points refer to granule cell nuclei, a whole point cloud to one subject. (**b**) Histograms of single nuclei feature plotted, example shown for one subject, revealing all features are approximately Gaussian distributed. (**d**) Violin plot of the heterogeneity for all subjects. (**d**) Median values of all subjects and each feature. A significant difference between controls and MS patients can be observed for the feature heterogeneity. Since *nn* takes only integer values, the mean instead of the median was used for reasons of accuracy. Note that all properties are shown centered to the population mean and normalized to the population standard deviation. (**e**) A matrix containing the Wasserstein-2 distance between any two individuals, calculated here between the histograms for the feature heterogeneity. Dashed lines separate the groups.

In fact, in addition to t-testing the median values, the entire distribution in the multidimensional feature space can be compared by metrics of optimal transport (OT) theory. Since the neuronal nuclei of each subject are not sufficiently well represented by only their median or mean values, comparing the entirety of recorded nuclei between subjects will allow for a more complete and powerful comparison, and hence also a more sensitive test of possible pathological alterations. Originally developed to model logistic transport problems, today OT is a popular tool in data analysis ***Peyré and Cuturi (2019***) since it allows measuring the similarity of distributions by the minimal “transportation cost”. As explained above, OT has the decisive advantage over classical statistical approaches that it takes the entire neuron population into account, enabling to detect small movements of subpopulations as well as to compare distributions not only in a single dimension (feature) but in high dimensional space taking all features into account simultaneously. We will use OT in two steps: first we compare single feature histograms one-by-one and then compare the full 6d point clouds with OT. To analyze the former, we compute the pairwise Wasserstein-2 distance between all histograms, which for two discrete measures *µ* and ν and Euclidean ground metric is formulated as

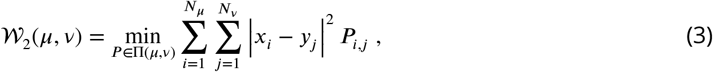

where **P** is the optimal coupling between *µ* and ν, Π the set of all couplings, and *x, y* denote the positions of the bins. For the calculation of 𝒲 the *Python Optimal Transport* - package by ***Flamary et al. (2021***) was used. The calculated pairwise Wasserstein distances are arranged in a matrix, which is shown in Fig. 4e. Dashed lines separate the two groups and divide the matrix into four quadrants. The higher values for 𝒲 in the upper right quadrant compared to the lower right and upper left quadrants implies that the distances between subjects of different groups are larger than within a group. This indicates a group segregation in the feature heterogeneity, as already suspected from the median values. For the remaining features, whose charts can be seen in the Appendix 3, again no clear trends can be identified.

### Multidimensional analysis with OT

In addition to the 1d-histograms, we next analyze the full six-dimensional point cloud distribution with optimal transport. Since the OT calculations of the point clouds are computationally expensive, they will be approximated by multidimensional Gaussian distributions, whose mean and covariance matrix are given by the empirical mean and covariance matrix of the point clouds. Note that the above analysis can also be applied to the point clouds, yielding the same results (compare Appendix 4). Figure 5a shows the Gaussian distributions in a 2d subspace represented by ellipses. The ellipses are centered around the mean, the orientation of the principal axes is given by the eigenbasis of the covariance matrix, and their length by the square root of the corresponding eigenvalues, which gives the 1-*σ* range around the mean. The Gaussians have the advantage that the Wasserstein distance between them can be calculated analytically by combining the Bures metric ***Forrester and Kieburg (2016***) on the covariance matrices Σ with the Euclidean distance on the mean values **m** according to

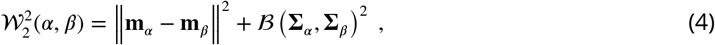

where the Bures metric ℬ is defined for positive definite matrices as

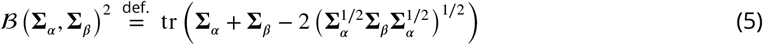

**Figure 5.**
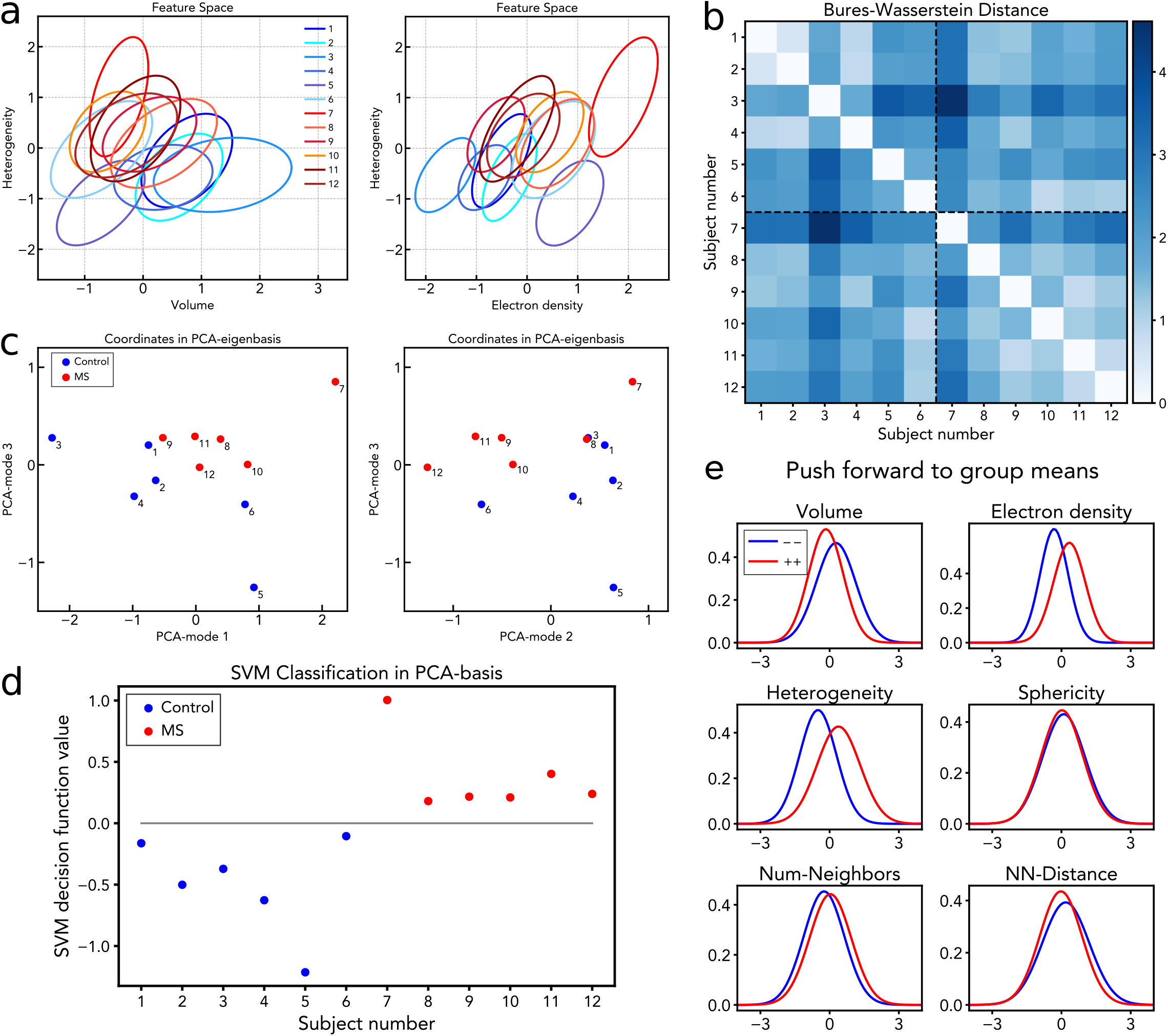
Optimal Transport Analysis. (**a**) The point clouds are approximated by multivariate Gaussian distributions, such that distances can be evaluated in closed form via the Bures metric (5)). The multivariate Gaussians are represented here by 1 − *σ* ellipses around the center of mass in 2d projections of the feature space. (**b**) After projection to the linear tangent space, all pairwise distances between the 6d ellipsoids are computed in linear approximation. Distances are arranged in a matrix chart, where dashed lines separate groups. (**c**) Following a PCA in tangent space, a low-dimensional embedding can be constructed, in which individuals are represented by single points instead of distributions. A segregation of the two groups into two clusters is observed. Note that this space is constructed without any prior knowledge about the sample classes. (**d**) Applying an SVM to the subject coordinates in the PCA-reduced space reveals a hyperplane separating the two groups. (**e**) The connection vector between the means of both groups in the subject space is used to generate “prototypical” distributions for both MS and Control via the so-called “push forward” operation. Histograms of these distributions, plotted for each feature and each group separately, inform about the pathological alterations of the nuclei from Control(- -) to MS(++).

Before using the Wasserstein distances in further steps below, it must be noted that the Wasserstein space of distributions is not a linear vector space. Contrarily, it forms a Riemannian manifold (curved hypersurface), which impedes straightforward application of standard linear algebra tools such as principal component analysis (PCA). To get around this, we follow the *Linearized Optimal Transport*-framework (LOT) introduced by ***Wang et al. (2013***) (for a review see ***Kolouri et al. (2017***)) in order to approximate the manifold locally by its tangent space at a suitable reference point (for full details see appendix 2 and references). As reference point, we choose the barycenter of all samples (whole population), which itself is a Gaussian distribution with mean and covariance matrix given by the fixed-point algorithm of ***Álvarez-Esteban et al. (2016***). After projecting the samples to the linear tangent space the Wasserstein distance between two embedded samples is approximated by the Euclidean distance between the embedding vectors. Figure 5 shows the linearized Wasserstein distances between any two subjects. The values are higher in the inter-group quadrant, than in the intra-group quadrants, (𝒲_inter_ = 2.039, compared to 𝒲_intraMS_ = 1.41, 𝒲_intraCTRL_ = 1.56) indicating a possible segregation of the groups. Moving to the tangent space comes with a change of perspective: in tangent space, each subject is now represented by a single point instead of a whole point cloud on feature space. We can thus interpret it as a “space of subjects”. The dimensionality of the tangent space equals that of the manifold of covariance matrices and mean values, given by 21 independent entries of the covariance matrix (accounting for symmetry) and 6 mean values, totaling in 27 dimensions. Before performing further analysis in tangent (or subject) space, we apply principal component analysis (PCA) to reduce the number of dimensions to 3. The three principal components capture 95% of the data variance. Plotting the coordinates of the subjects in the reduced 3d PCA eigenbasis as depicted in Fig. 5c, reveals that the subjects form two clusters according to their groups. Each principal component contributes almost equally to the segregation of the groups. Note that the construction of the subject-space is done without any prior categorization into groups. We further apply a simple linear support vector machine (SVM), in 3d which returns a hyperplane for classification. Figure 5d shows the distances of the samples to the hyperplane. The hyperplane divides the samples exactly into their classes, demonstrating the data are linearly separable. By the so-called “push-forward” it is possible to map from the tangent space back to the space of Gaussian distributions and thus movements in tangent space can be translated to changes in the distributions of the individual features. We use this to study the difference between the prototypical distributions of both classes. For this purpose, the mean values of both groups in the subject space are calculated, whose difference vector can be interpreted as the main direction of discrimination between healthy and pathological. Via the “push-forward” we then generate distributions corresponding to the (purely virtual) subjects obtained by moving from the origin of the tangent space along the difference vector, once into the Control- and once into the MS direction. We interpret these as prototypical representatives of MS- and Control subjects. From these distributions, histograms can be generated for each feature separately, which are shown in Figure 5e. It can be seen that the histograms differ in several features. This result allows to indicate a pathological transformation, which the granule cell nuclei undergo during the disease. According to the histograms, granule cells of multiple sclerosis patients compared to those of healthy Controls have:

- a smaller volume *ν*
- a higher electron density *ρ*
- a higher heterogeneity *s*

## Discussion

The first structural property and question to be discussed is a simple one: does the spatial density of neurons, which is already exceptionally high in the granular layer of the cerebellum, increase further in MS, and if so why could this possibly the case? Indeed, this study finds a 16% increase in density for the MS group, albeit only at marginally statistical significance *p* = 0.09. Cell segmentation and counting in the reconstructed volumes gave a direct assessment of cellular density, obviously superior to conventional estimates based on 2d observation. The increase is further corroborated and detailed by comparing the structure factors *S*(*q*) describing the short range order of granule cells. Note that the center of mass positions for all cellular nuclei in a certain volume makes it possible to statistically analyze the short range order in quantitative terms. Here, the overall increase in density is reflected by a shift of the first maximum of *S*(*q*) towards higher *q*, indicating a smaller next neighbor distance in the MS group. We can tentatively put forward the following interpretation: Since it is unreasonable to assume that new neurons have been formed in the course of the disease, the observation of higher density and shorter next neighbor distance could be explained only by tissue shrinkage, possibly as a response to a less active state of neurons (see below) and tissue remodeling in the inter-neuronal space, the neuropil. This would be in line with earlier studies which discussed tissue loss and brain atrophy as a result of axonal damage by demyelination and neurodegeneration ***Weier et al. (2015***); ***Lassmann et al. (2007***); ***Haider et al. (2016***).

Next, we address the structural properties (features) of the neuronal nuclei: heterogeneity, which quantifies the density variation with the nucleus, is the most significant feature changing between MS and Control. Already on the level of the median values it shows a significant increase. This if further corroborated by OT analysis, which compares the entire histogram of a feature, and therefore can also account for changes in neuronal population of a subject when its mean value remains constant. If, for example, transitions occur in the width or shape of the distribution such as increased tails, this may not affect median or mean by left/right symmetry but clearly changes the distribution. In this sense, OT is a more complete and more sensitive probe of structural alterations between the groups. For a more transparent analysis we approximate the non-linear OT space by a linear tangent space and subsequent reduction with PCA to three dimensions. The resulting embedding shows a clear separation between the groups, which is identified without a prior hypothesis by the OT analysis. A simple SVM classifier is then able to perfectly separate the two classes along an axis. Alternatively we consider the axis spanned by the difference of the class means. Via the push-forward we find that this axis encodes a transition to increased heterogeneity, smaller volume, and higher density. We might call it an axis of compactness.

How can this shift towards a more compact nuclear state, i.e. more heterogeneous, smaller and denser nuclei be interpreted? By considering the spatial scales which contribute to heterogeneity, it is plausible to attribute this feature to an increased ratio of heterochromatin to euchromatin ***Le Gros et al. (2016***). In fact, we put this interpretation forward in our preceding study on hippocampal granule cells in AD, where a very similar observation was made ***Eckermann et al. (2021***).

Accordingly, a transcriptionally less active state of the neuron corresponds to the more compactified nucleus. These states could be interpreted as a phenomenon of cellular senescence ***Kritsilis et al. (2018***). The fact that this is found similarly in the present work for cerebellar granule cells in MS as for hippocampal granule cells in AD before, suggests the hypothesis that a more compact nucleus resulting from cellular senescence is a more general phenomenon in neurodegeneration, downstream from various patho-metabolic processes.

The largest weakness of the present study is its still too small size (*N*_*MS*_ = 6/*N*_*CT RL*_ = 6). The power of OT may not or not sufficiently compensate for this, and a higher number of subjects should certainly be probed in future extension of this work. While this would not be much of a problem per se in view of method throughput (XPCT data acquisition and analysis with a fully automated pathway), the post mortem collection of human tissue is not easily extended to higher numbers, given necessary procedures of consent and authorizations. Further improvements may require concerted efforts in operation of tissue banks, proper documentation, and curated collections.

What the current study has not touched upon is the important role of demyelinated lesions in MS. We did not make any attempt to find specific structural signs of lesions or to identify them by correlative imaging with immunohistochemistry. To this end, one must also critically put into question whether this is best carried out with unstained and unlabeled tissue as in the present case, or whether heavy metal stains or labels for XPCT would be required, for example also to locate regions of de- and remyelination. Further, it would make sense to increase the scan volume at the cost of lower resolution, and to use very clear cases to ‘train’ any search for lesions. Given the substantial role that magnetic resonance imaging (MRI) can play in MS diagnosis as an in vivo imaging method ***Wattjes et al. (2015***), there is a further very worthwhile goal for future extension of this work: using XPCT analyses and correlative XPCT/MRI imaging, one could correlate the post mortem 3d histology and the tissue fine structure with more coarse grained but also functional signals of MRI. Note that as 3d imaging technique XPCT is particularly well suited for a multiscale histopathology underpinning of MRI data.

Finally, a critical reflection regarding the relevance of structural data: while it is undisputed that genomics, proteomics and metabolics are relevant to gain a quantitative understanding of neurodegenerative diseases, the relevance of the cytoarchitecture is admittedly less clear. In view of the intrinsic polydispersity of structural features on the cellular and tissue level, differences between individuals can easily screen effects associated with disease progression. Further, it is less known than for biochemical processes, whether structural alterations are upstream or downstream from a particular pathological development. If studies of cytoarchitecture are to become an ‘omics’, structural data has to be very comprehensive, covering large patient- and control-groups, quantitative and fully digital. It should certainly also represent the full three-dimensionality of tissue. Finally, without segmentation and morphometric analysis, 3d data alone will remain illustrative and anecdotal, since visual inspection is not as easily possible as for 2D sections by a pathologist. While the present work can surely not meet all expectations of how structural brain tissue studies should be carried out in future, it is meant as an example and to provide useful components to further develop the analysis workflow. We can expect significant future progress in segmentation by deep learning and in optimal transport theory, as for the data acquisition itself and its image quality.

In order to help this become a reality, the present work is carried out as part of a larger effort to advance quantitative assessment of neuronal cytoarchitecture by XPCT. The method can extend conventional histology by a further dimension and therefore is particularly well suited for digitalization and automated analysis of tissue structures. To this end it is a decisive advantage that XPCT does not rely on tissue sectioning, is non-destructive, and compatible with all other analyses which can be carried out subsequently. Furthermore, XPCT can be performed on unstained tissue preserved in FFPE blocks, which is the conventional way to store and preserve tissues in neuropathology.

## Materials and Methods

### Data acquisition and reconstruction

Tissue asservation, data acquisition, phase retrieval and tomographic reconstruction of all data analyzed here was performed previously, as reported in ***Töpperwien et al. (2018***). In short, human cerebellum tissue samples were obtained post mortem from twelve individuals (six healthy control, six multiple sclerosis) by routine autopsy in agreement with local ethics guidelines and approval procedures at the University Medical Center Göttingen. Small biopsy-punches from the formalin fixed and paraffin embedded tissue (FFPE) were placed in a Kapton tube for scanning. X-ray phase contrast tomography experiments were carried out at the GINIX endstation of the P10 undulator beamline at the PETRAIII storage ring at the Deutsches Elektronen Synchrotron (DESY) in Hamburg. The undulator beam was monochromatized to an energy of 13.8 keV (Si(111) monochromator). Note that for one sample (CTRL5, different beamtime), the data was collected at 8 keV. After prefocussing the x-rays by a pair of Kirkpatrick-Baez (KB) mirrors and coupling into a waveguide, the coherence and spatially filtered beam illuminates the sample at distance *z*_01_ *≃* 0.1 m behind the waveguide exit, and the magnified Fresnel diffraction pattern (hologram) is recorded by a fibre-coupled sCMOS detector positioned at distance *z*_02_ *≃* 5.1 mm, resulting in a geometric magnification 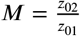. From the measured magnified holograms (wave optical projection images), phase retrieval was performed using the contrast transfer function (CTF)-based algorithm ***Cloetens et al***.(***1999***), implemented in ***Lohse et al. (2020***). The 3d information was reconstructed with the Matlab implemented function of the inverse Radon-transformation (‘iradon’) combined with a standard Ram-Lak filter. The reconstructed samples cover a field of view of 336 × 336 × 375 µm^3^ with a voxel size of 187 nm, sufficient to resolve various histological features, including the nuclei of the granule cells.

### Segmentation of the granule cell nuclei

The segmentation was carried out with the segmentation and visualization software package Arivis Vision4D (Zeiss AG, Germany). Using the Blob Finder operation of Arivis - well suited to find round, roughly spherically shaped objects - several ten thousand neurons were detected in each sample. After applying different filters and removing objects outside the granular layer with a mask, a homogeneous distribution of sphere-like segments was obtained, adequately representing the granule cell nuclei. From the segmented nuclei, several features were extracted for the analysis. Detailed information about the full segmentation workflow is given in Appendix 1.

### Structural features of the granule cell nuclei

For the analysis, six features of the segmented nuclei were chosen: the volume *ν*, the mean of the electron density *ρ*, the heterogeneity (variance of the electron density within the nucleus) *s*, the sphericity *ϕ*, the distance to the nearest neighbor nuclei *d*_*nn*_ and the number of neighbors *nn* within a radius of 7.45 µm. The radius for the latter definition was chosen as the local minimum between the first and second correlation shell of the pair correlation function *g*(*r*).

### Optimal Transport analysis

Optimal transport distances between 1d feature histograms were computed by using the Wasserstein - 2 metric 𝒲_2_, as implemented in ***Flamary et al. (2021***). For the analysis of the multidimensional distributions, each point cloud was approximated by a normal distribution with covariance matrix Σ and mean *µ*. Between Gaussians, the Wasserstein metric can be rapidly calculated by using the Bures metric ℬ, see ***Forrester and Kieburg (2016***). To overcome the Riemannian structure of 𝒲, we used the *Linearized OT* framework as described in ***Wang et al. (2013***) and project the distributions into a linear tangent space. An approximate Wasserstein barycenter, computed by the fixed-point algorithm ***Álvarez-Esteban et al. (2016***), served as reference point for linearization. This allowed us to construct a subject space, in which subjects were arranged in two clusters according to their groups (without prior classification) and could be linearly separated by SVM (implemented in ***Pedregosa et al. (2011***)). For more details of the OT framework, see Appendix 2.

## Data Availability

All data produced in the present study are available upon reasonable request to the authors.

## Acknowledgments

We thank Jannis Schaeper for initial help with the Arivis software. T.S. and B.S. acknowledge support by the Deutsche Forschungsgemeinschaft (DFG, German Research Foundation) through project CRC 1456/ A03. We also thank Markus Osterhoff for IT support in the framework of the data infrastructure project SFB 1456/ INF. C.S. received funding from the Deutsche Forschungsgemeinschaft (DFG, German Research Foundation) – CRC 274/1-Project ID 408885537 B01, the DFG Sta 1389/5-1 (individual research grant). C.S. and T.S.are supported by the DFG under Germany’s Excellence Strategy (EXC 2067/1-390729940). J.F. was supported by the clinician scientist program of the CRC 274.

## Appendix 1

### Segmentation Workflow

**Appendix 1 Figure 1.**
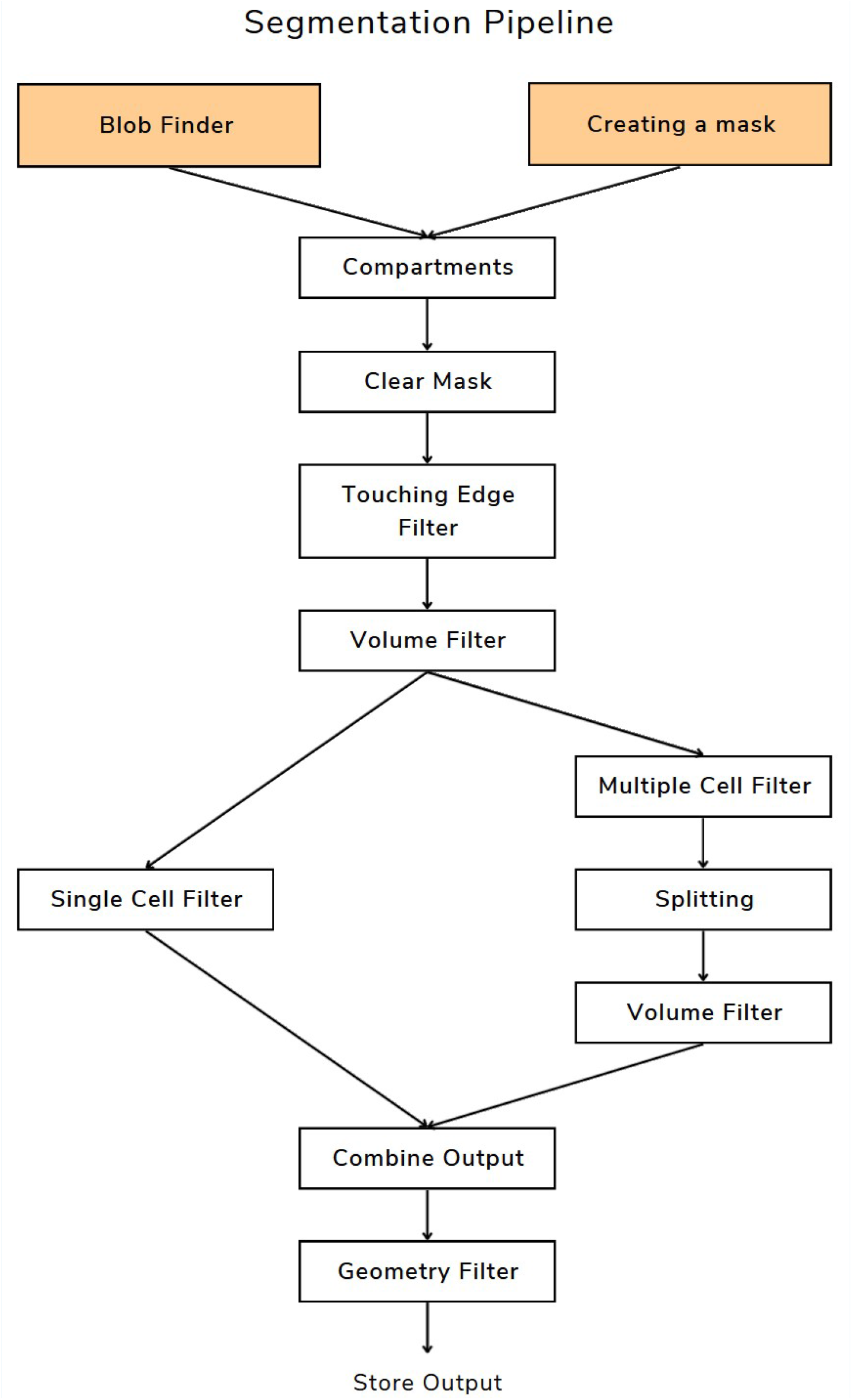
Flowchart of the Arivis pipeline used for the segmentation of the granule cell nuclei. Orange fields indicate segment generating operations.

In order to segment the granule cell nuclei, the software Arivis was used. The workflow in Arivis is performed by pipelines - a sequence of operations which are executed consecutively. In the following the structure of the pipeline used for the segmentation is presented and the individual operations are briefly described.

- **Blob Finder:** The Blob Finder is the most important operation which generates the segments representing the granule cell nuclei used in the analysis. The algorithm is designed to find rounded, sphere-like 2d or 3d objects and has three parameters which can be customized. The concept is to find seed points with high probability to be the centre of a round object, followed by a watershed algorithm, letting objects grow outwards starting from the seed points. The following description of the algorithm is based on the Arivis manuals. Consider a gray scale image *f* : ℝ^3^ → ℝ which is convolved by a Gaussian Kernel *g* with:

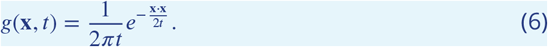

**x** represents the pixel position and *t* is the scale of the convolution kernel given by

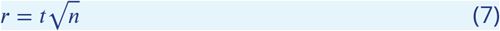

where *n* is the dimension (here = 3) and *r* is the “diameter”, which is one of the parameters that can be set manually. After the convolution

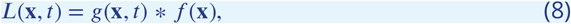

a Laplace operator is applied to the result ∇^2^*L*(**x**, *t*) which gives a probability map of possible blobs. This probability map is thresholded to create a binary mask and to find the seed points.

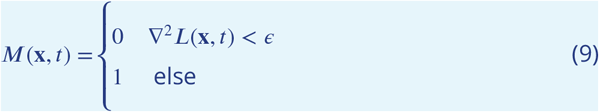 The adjustable parameter *ϵ* is used to vary the number of considered objects. Subsequently, the local maxima of the Laplacian image ∇^2^*L*(**x**, *t*) are used as markers for a topological Watershed transform ***Najman and Couprie (2003***). The result is masked with *M*(**x**, *t*). In this way *ϵ* determines not only the number of created objects, but also their size. The last parameter is the “split sensitivity”, which defines whether two seed points in close vicinity are merged together or remain separated. The parameter operates like a threshold on the values of the probability map and determines whether many small objects or a few larger objects are observed. For the segmentation of the GC nuclei, the parameter were set as follows:
  – Diameter: 3.74 µm
  – Probability Threshold: 26 %
  – Split Sensitivity: 24.72 % These parameters were determined by visual inspection and were chosen the same for all samples.
- **Creating a mask:** Next a mask is drawn manually, which encloses only the granular layer. Segments created by the Blob Finder lying in the molecular layer, Purkinje layer or in the corners of the dataset, where the tomographic reconstruction does not yield completed information, are filtered out.
- **Compartments:** This operation creates a hierarchy between the mask and the Blob Finder segments. All segments which are not part of this hierarchy, i.e. all segments outside the mask, will be filtered out.
- **Clear mask:** Removes the mask object.
- **Touching Edge Filter:** This filter removes segments touching the edges of the dataset, since these could be truncated cells which would distort the statistics.
- **Volume Filter:** The volume filter removes segments which clearly do not represent granule cells, e.g. small artifact segments. The filter limits are determined by displaying all segments in a 2d scatter plot according to the properties volume *ν* and sphericity *φ* (using the Object chart tool of Arivis) as shown in figure 2b. In this representation, three groups of segments can be identified: granule cells (middle), multiple cells (middle) and artifacts (right). This assertion is corroborated by selecting segments of each group and inspect the 3d rendering. The artifact segments are filtered out, and the multiple cell segments are kept and split (see below). By visual inspection of the scatter plot, the filter limits are set to *ν* < 10 µm^3^ and *ν* > 100 µm^3^.
- **Multiple Cell Filter:** Multiple cells occur when several cells close to each other are covered by only one segment. To ensure each cell is covered by exactly one segment, the multiple cell segments can be split (next operation). By visual inspection of the scatter plot, filters of *φ* < 0.58 and *ν* < 30 µm are determined to segregate the multiple cells from the single cells.
- **Splitting:** The splitting operation divides the multiple cell segments into well-defined segments covering single cells. The splitting uses a Watershed-Algorithm which takes the maxima found in a distance map as seed points. The distance map labels each pixel of a segment with the distance to the nearest non-segment pixel. The operation has the parameter “split sensitivity”, which operates like a threshold in the watershed algorithm and controls into how many parts the segments are split. For all samples, a value for the split sensitivity of 70% was chosen.
- **Volume Filter:** After splitting, small artifact segments can occur which can be removed again with a volume filter of *ν* > 10 µm^3^.
- **Combine Output** The single cells and the split multiple cell are merged to one group representing the granule cells.
- **Geometry Filter:** Finally, segments are removed which have an elongated, non-round shape. They can occur due to the segmentation of blood vessels and other tissue structures. To discriminate them from the granule cell segments, a bounding box in the shape of a cuboid is calculated (Arivis function) for each segment. Segments where the long side of the bounding box being 2.5 times larger than the shortest side are removed.

Note that the limits for the different filters may slightly vary from sample to sample, but always remain very close to the values specified above. From the segmented granule cells, the following properties are extracted and stored in .txt-tables: Center of geometry in x, y, z-position, volume, sphericity, mean value of intensity (gray values), standard deviation of intensity (gray values).

## Appendix 2

### Optimal Transport Analysis

**Appendix 2 Figure 1.**
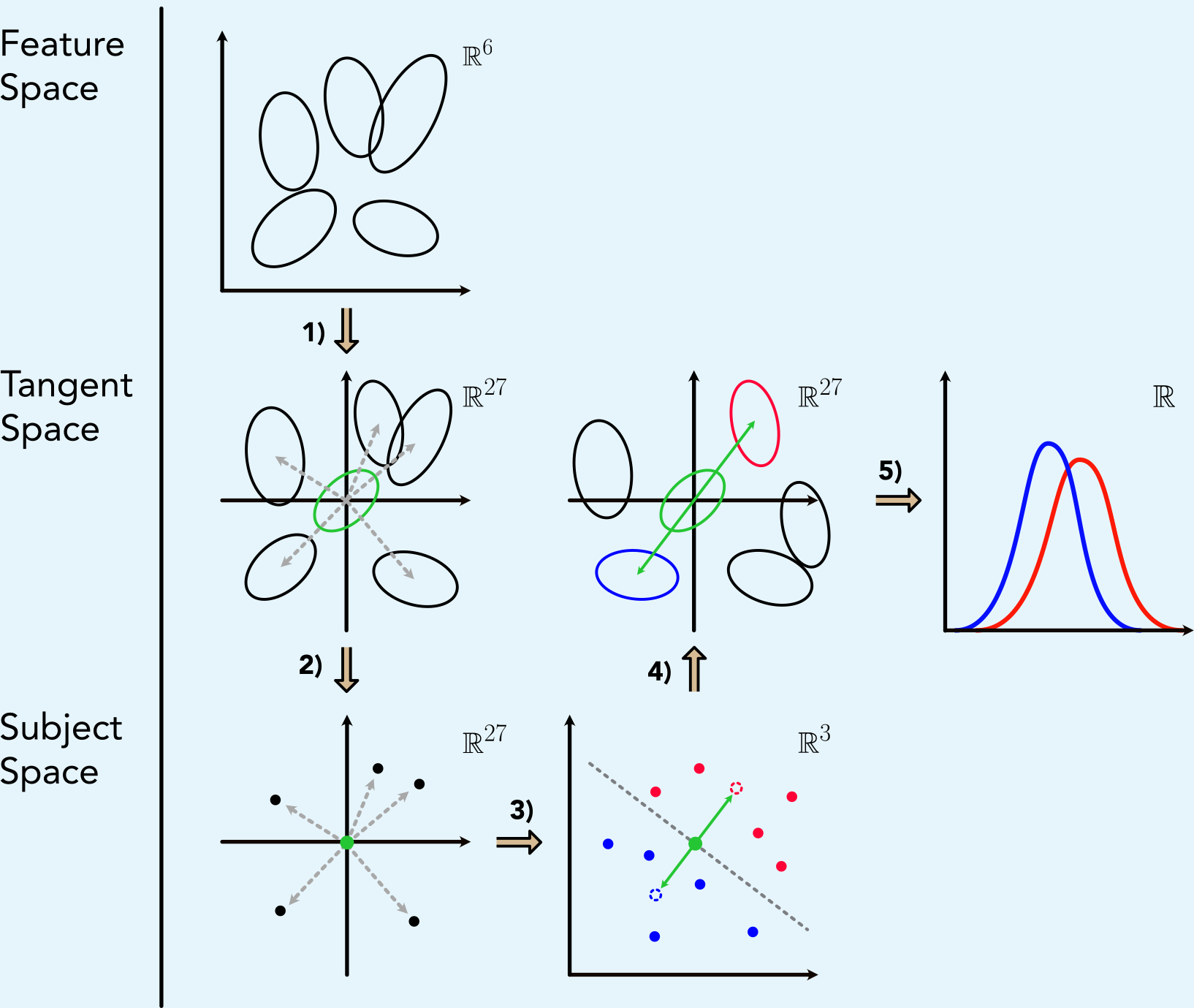
The figure schematically illustrates all steps of the optimal transport workflow starting from the Gaussian representation of the subjects to the final so-called push forward histograms. The individual operations shown in the scheme here are described in detail below.

For the multidimensional analysis with OT, we used the *Linearized Optimal Transport* - framework ***Wang et al. (2013***) which provides several advantages. The key idea is to pick a reference sample and calculate the pairwise optimal transport plans and Wasserstein distances between the samples and the reference sample instead of calculating them between all samples. All other pairwise distances can then be in a simple way approximated from the initial *M* optimal plans. Hence, one needs to compute for *M* samples only *M* plans rather than *M*(*M* − 1)/2 distances. This also implies a linear approximation of the Riemannian structure of the Wasserstein space (formally a curved hypersurface), which enables to apply standard analysis tools. In the following, it is described by the figure above, how we applied the framework to the Gaussian data. The described steps correspond to that in the figure.

- **1) Local Linearization:** Consider a point cloud distribution for each individual, obtained by locating all GC nuclei according to their features. In a first step, we approximate the point clouds by Gaussian distribution to strongly reduce the computational effort. This is valid since each single feature is well approximated by a Gaussian distribution. The Gaussians are determined by the empirical mean and covariance matrices of the point clouds. Following the LOT framework, we first compute the Wasserstein barycenter *σ* which serves as reference point for the local linearization (green ellipse). The barycenter itself is a Gaussian distribution with mean and covariance matrix approximately calculated by the fixed point algorithm ***Álvarez-Esteban et al. (2016***). Starting from the barycenter, each subject can now be projected into the linear tangent space via the Riemannian logarithmic map, in this case given by

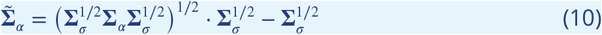

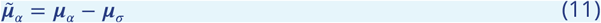

where **Σ**_*σ*_ is the covariance matrix of the barycenter and **Σ**_*α*_ the covariance matrix of the corresponding sample. These can be interpreted as encoding the direction from *σ* to *α*, visualized in the figure by grey arrows representing the linear embedded samples.
- **2) Interpretation as Subject Space:** After projecting the samples in the tangent space, we take on a different perspective. For each sample we now consider the combined and flattened entries of 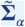 and 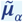as a single vector. Thus, a single point / single vector *x*_*n*_ with 27 dimensions (21 independent covariance matrix entries, accounting for symmetry, and 6 mean values) is obtained for each subject. This space is denoted as the subject space, in which subjects are represented as single points and not by Gaussians anymore, but information about mean and covariance of the samples are preserved. The pairwise Wasserstein distances between subjects can now be simply approximated by the euclidean norm on the coordinates. **3) PCA and SVM** To better investigate the high dimensional subject space, we further reduce its dimensionality by a principal component analysis (PCA) and truncation of the number of dimension from 27 to 3. This choice is based on the spectrum of eigenvalues, where the first 3 eigenvalues cover 95% of the variance while the rest does not significantly contribute. Solving the eigenvalue problem and arranging the eigenvectors to a transformation matrix **T**, the projection of the samples ***x***_*n*_ (after centering) to the eigenbasis is carried out according to:

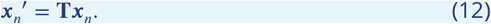

In this representation, the subjects fall - without any prior information - into two clusters corresponding to the two groups. We perform a simple classification approach by applying a linear support vector machine (SVM, regularization parameter *C* = 5) to the subjects in the reduced 3d PCA-eigenbasis. The SVM returns a hyperplane exactly segregating the subjects according to their groups. The obtained normal vector of the hyperplane is used to calculate the distances of the samples to the plane (see Fig. 5d). Next we investigate how this clear discrepancy between MS- and Control group is reflected in the individual features. To do so, the mean points of both groups are calculated in the 3d subject space, whose difference vector **N**′ can be considered as an alternative axis of discrimination (which we expect to be more robust than the axis given by SVM). Moving along this axis to the group means, results in two virtual points representing a typical MS- and a typical Control subject (blue and red dashed circle).
- **4) Back Projection to Gaussian Distributions:** From the points in the 3d PCA - eigen-basis, Gaussian distributions with covariance matrix and mean can now be generated by inverting the above transformations. First, we return from the reduced eigenbasis to the full tangent space ℝ^27^ using the inverse of the transformation matrix *T* ^−1^

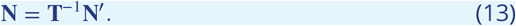

Afterwards the vectors in ℝ^27^ can be decomposed back to the covariance matrix and mean, according to the inverse order as in the initial transformation. This way a Gaussian distribution is obtained for the typical MS- and Control sample which can be compared (blue and red ellipses). This corresponds to the push-foward of the barycenter under the respective tangent vectors.
- **5) Push forward histograms:** Based on the Gaussians for the typical Control and MS patient, histograms can be generated for each of the six features separately and compared. The resulting histograms, shown in Fig. 5e, reveal the pathological alteration of the granule cells towards a more compact structure in MS.

Note that other axes can also be selected as push forward direction **N**′, such as the normal vector of the SVM hyperplane or the PCA principal axes (see also ***Eckermann et al. (2021***)). Further, we evaluated the data also on the level of point clouds. The workflow is completely analogous to the Gaussians with a few exceptions: The reference sample *σ* is (analogous to the Gaussians) approximated by the fixed-point algorithm but from the resulting ***µ*** and **Σ** we sample a point cloud distribution with 10^4^ particles and uniform weights

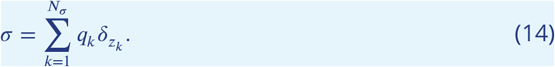

Local linearization is then performed by solving the transport problem between samples and reference sample with entropic regularization and Sinkhorn’s algorithm ***Cuturi (2013***). From the resulting couplings, we calculate mean mass transport from the particles of the barycenter *z*_*k*_ to each sample. The connections between *z*_*k*_ and the centers of the averaged transport now become the set of approximated tangent vectors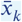. Thus, we obtain an approximated coupling Π induced by a Monge map. Between two samples *η* and ν, the Wasserstein distance can then be approximated by the ℒ^2^ norm on the set of tangent vectors

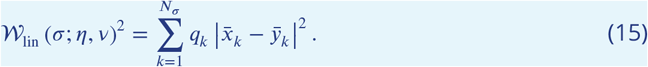

## Appendix 3

### Supplementary Plots

**Appendix 3 Figure 1.**
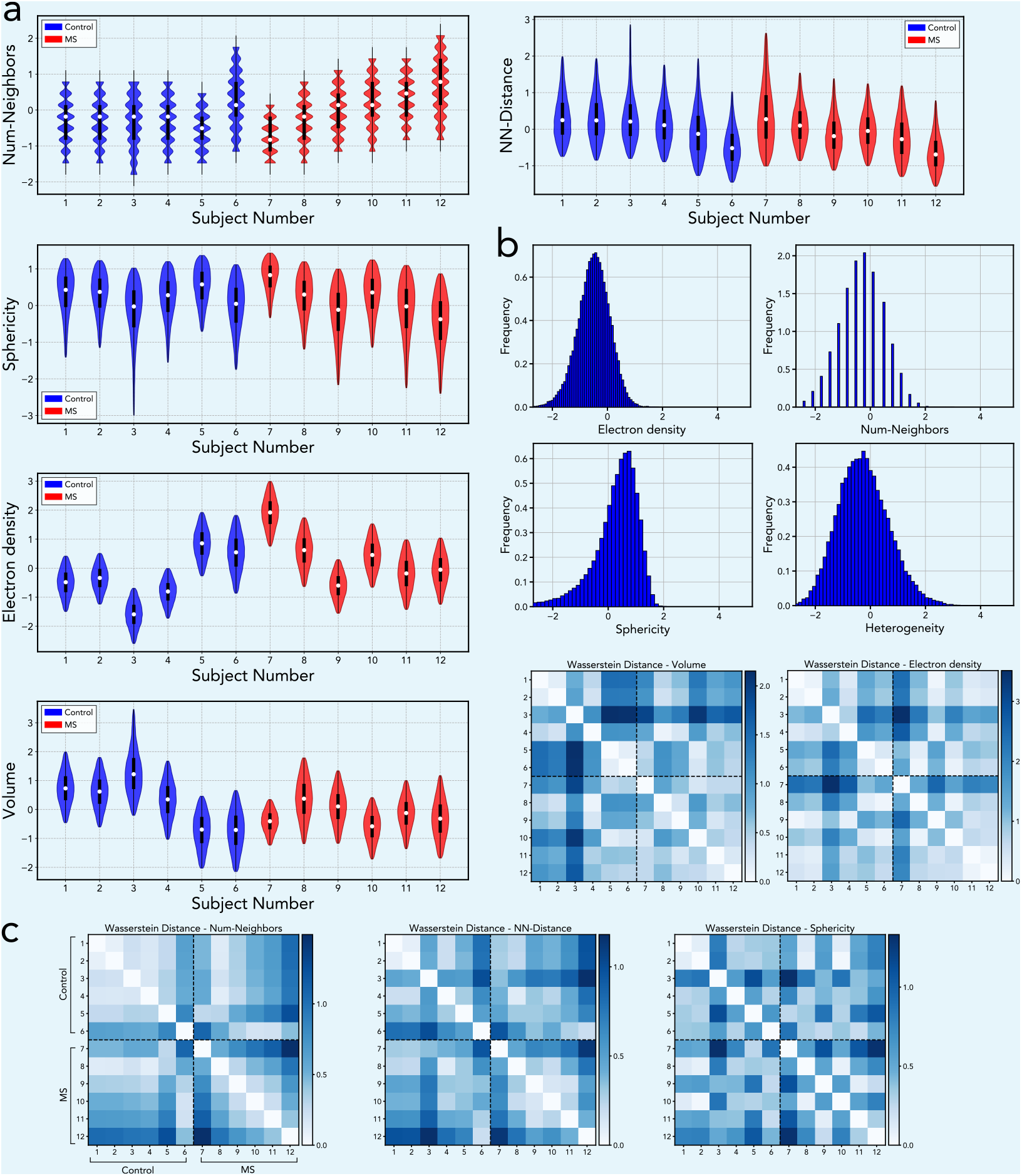
The figure shows all supplementary plots of the 1d analysis, which were not shown in the main part. (**a**) Violin plots of structural features for each individual. Note that the wavelike structure of the number of neighbors - plot is due to the fact that this feature always takes on integer values. (**b**) Histograms of structural features revealing all features are approximately Gaussian distributed. (**c**) Matrix chart of pairwise Wasserstein-2 distances between 1d feature histograms of all individuals.

## Appendix 4

### Supplementary Plots

**Appendix 4 Figure 1.**
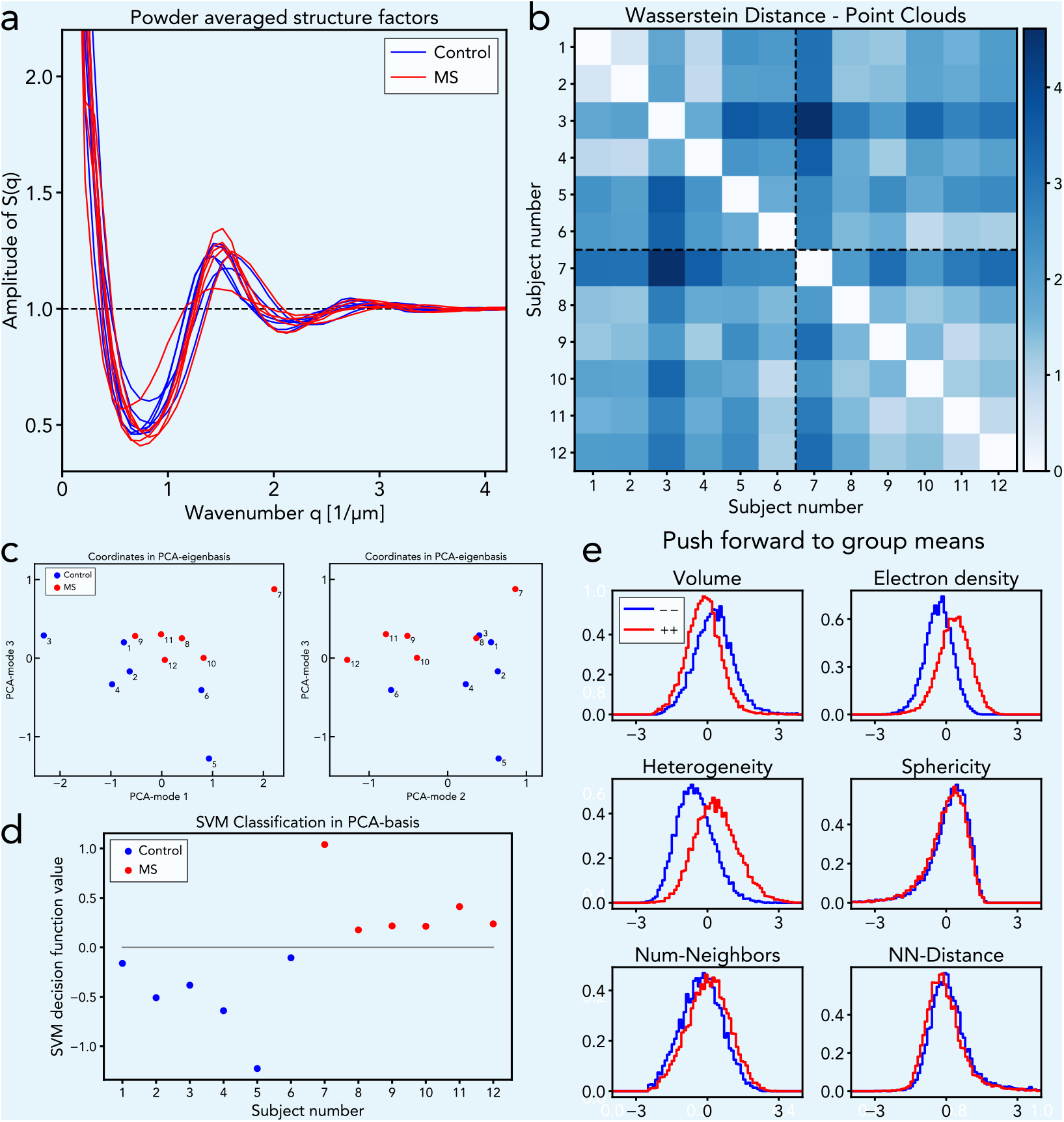
Structure factors and OT-analysis of the point cloud distributions (**a**) Powder averaged structure factors of each subject. By averaging over both group, the structure factors shown in Figure 3a are obtained. (**b-e**) OT analysis of the point cloud distributions with a workflow mostly analogous to the Gaussian distributions. The results for the Wasserstein distance chart (b), the subject space in PCA-eigenbasis (c) and the SVM classification (d) are very similar to those obtained in the analysis based on the Gaussian approximation. In the push forward of the barycenter (e), one obtains a histogram of a point cloud rather than that of an Gaussian. However, the trends of the individual features, towards compact nuclei in MS, are the same. Overall, the Gaussian distributions can be considered as a valid approximation of the point clouds, giving very similar results at a very low computational cost.

